# Egocentric Sexual Networks of Men Who Have Sex with Men in the United States: Results from the ARTnet Study

**DOI:** 10.1101/19010579

**Authors:** Kevin M. Weiss, Steven M. Goodreau, Martina Morris, Pragati Prasad, Ramya Ramaraju, Travis Sanchez, Samuel M. Jenness

## Abstract

In this paper, we present an overview and descriptive results from the first egocentric network study of MSM from across the United States (U.S.): the ARTnet study. ARTnet was designed to support prevention research for human immunodeficiency virus (HIV) and other sexually transmitted infections (STIs) that are transmitted across partnership networks. ARTnet implemented a population-based egocentric network study design that sampled egos from the target population and asked them to report on the number, attributes, and timing of their sexual partnerships. Such data provide the foundation needed for estimating and simulating stochastic network models that are used for disease projection and intervention planning. ARTnet collected data online from 2017 to 2019, with a final sample of 4904 participants who reported on 16198 sexual partnerships. The analytic aims of the study were to characterize the joint distribution of three network parameters needed for modeling: degree distributions, assortative mixing, and partnership length, with heterogeneity by partnership type (main, casual and one-time), demography, and geography. Participants had an average of 1.19 currently active partnerships (“mean degree”), which was higher for casual partnerships (0.74) than main partnerships (0.45). The mean rate of one-time partnership acquisition was 0.16 per week (8.5 partners per year). Main partnerships lasted 272.5 weeks on average, while casual partnerships lasted 133.0 weeks. There was strong but heterogenous assortative mixing by race/ethnicity for all groups. The mean absolute age difference was 9.5 years, with main partners differing by 6.3 years compared to 10.8 years for casual partners. Our analysis suggests that MSM may be at sustained risk for HIV/STI acquisition and transmission through high network degree of sexual partnerships. The ARTnet network study provides a robust and reproducible foundation for understanding the dynamics of HIV/STI epidemiology among U.S. MSM and supporting the implementation science that seeks to address persistent challenges in HIV/STI prevention.

## INTRODUCTION

Human immunodeficiency virus (HIV) and other sexually transmitted infections (STIs) continue to present significant public health challenges. In the United States, HIV and STI incidence disparities are linked to demographics (1), risk behavior (2), clinical care access (3), and geography (4). Of the estimated 40,000 new HIV infections occurring in 2017, two-thirds were among men who have sex with men (MSM) (5). The large disparities in HIV/STI cases by race and age have worsened, with incidence increasing among younger non-white MSM while decreasing in other MSM groups (6). Syphilis has also concentrated among MSM (7), following similar demographic and geographic patterns as HIV (8,9). Understanding the persistent and emerging drivers of HIV/STI transmission dynamics among MSM is critical to prevention.

Sexual partnership networks are the mechanism through which all STI and most HIV transmissions circulate. The pathogens are transmitted by sexual acts embedded within partnerships, and circulation through the population depends on how those partnerships form and dissolve — a highly structured and population-specific dynamic process (10–12). While sexual network structure can be measured and analyzed either cross-sectionally or dynamically (13), dynamic networks determine how infectious diseases spread when the average duration of infection is longer than the average duration of partnerships (14). This is often the case for HIV/STIs in the context of persistent sexual partnerships.

A large body of research over the past three decades has investigated which network features are most important in determining the size and speed of HIV/STI epidemics within and across populations (15). Three key features are the distribution of degree (number of ongoing partners), assortative mixing (selecting partners based on attributes similar to one’s own), and the distribution of partnership durations (14). Partnership concurrency (2+ ongoing partners) amplifies the speed of HIV/STI transmission by allowing for backward paths of transmission (from infected partners acquired later to earlier uninfected partners) and shorter waiting times before onward transmission (10). Heterogeneity in network degree and duration, paired with assortative mixing by demographic attributes, may also play an important role in generating persistent disparities in HIV/STI incidence between MSM subgroups (16,17). Network definitions also implicitly underlie recommendations for HIV/STI prevention tools, including guidelines for HIV preexposure prophylaxis (PrEP) that outline indications based on network degree, partnership type, and mixing by disease status (18).

Recent methodological advances have removed the primary obstacle to network measurement for HIV/STI epidemiology: the absence of a general statistical framework for temporal network model estimation from sampled network data (19). Traditional descriptive network analysis has relied on the collection of data on a complete cross-sectional network census — measurement of all nodes (persons) and edges (partnerships) in a population. While this design has been attempted in a couple of high-profile sexual network studies (20,21), cost and implementation challenges make it infeasible for routine public health use. Various forms of adaptive sampling have also been developed, of which Respondent-Driven Sampling (22) and partial sociometric designs (23) are most well-known. However, methods for recovering valid estimates of the population network structure from these designs are limited and do not include temporal networks needed for disease modeling (24). The methodological advance that changed the field was principled estimation of Exponential-family Random Graph Models and their temporal counterparts from egocentrically sampled network data (25). Egocentric network sampling draws egos (study participants) from the population of interest and then asks them to report on the number, attributes, and timing of their partnerships (12). It requires no link tracing, and can be integrated into a standard survey sampling design. Statistical methods can then be used to infer the complete dynamic network structure consistent with the sampled network data.

Temporal exponential random graph models (TERGMs) are an inferential statistical framework for complete or egocentrically sampled network data. They can be used to estimate the population parameters of network models from egocentric sample statistics on the incidence, duration, and heterogeneity in sexual partnerships at the dyadic (partnership) level (19). Once estimated, TERGMs can then be used for simulating from the temporal network defined by the model. The simulated networks will reproduce, on average, the observed (or scaled) network statistics from the sample. This provides an empirically grounded approach to simulating the partnership networks needed for mathematical projection models of infectious disease dynamics (25). Software such as EpiModel provides flexible tools for building network-based epidemic models with egocentrically sampled network data (14).

Egocentric data provide directly observed information on a limited but key set of network structural features: degree distributions, assortative mixing by nodal attributes, and partnership duration, all of which can be stratified by demographic and partnership attributes. However, not all structural features can be observed in this design. Unobserved features include assortative mixing by degree (that goes beyond mixing by measurable attributes) and higher-order aggregate network features, like geodesics and component size distributions, not measurable one or two edges removed from the sampled ego. However, many of these aggregate network features are largely determined by the lower order features that *are* observable — they are emergent properties of the local rules people use for partner selection — so the loss of information is less than it might appear.

Egocentric data have been successfully used to model the drivers of HIV transmission, racial disparities in disease incidence, and optimal strategies for HIV/STI prevention (26–28). These empirically grounded network models depend on egocentric network data. The Demographic and Health Surveys have been a useful data source for populations in Sub-Saharan Africa (24), but to date there have been no network data sources for MSM (in the United States or elsewhere) that broadly span across demographics and geography. MSM network data commonly used in epidemic models (including our own) have been collected in cohort studies with restrictive eligibility criteria (17), aggregated across multiple small studies in ways that may impact inference (29), or transported across studies of populations not represented in the models (30). The HIV/STI modeling field needs up-to-date estimates of commonly used network parameters from study designs without restricted behavioral eligibility criteria for parameterizing network models for HIV/STI at multiple scales and in multiple geographic settings.

In this paper, we present descriptive results from the ARTnet study, the first egocentric network study of MSM from across the United States. Our analytic aims were to characterize the distribution and heterogeneity in network degree, assortative mixing, and partnership length (a proxy of partnership duration useful for modeling). We estimated these three outcomes with stratifications by geography, demographics, and HIV status for different sexual partnership types (main, casual, and one-time) and different definitions of what type of sexual intercourse constitutes a sexual partnership (anal versus oral intercourse). The results in this paper provide summary network statistics that can be used in future epidemic modeling applications. We will also be making the full data available for primary analysis by external researchers, so these results represent just a small selection of the potential model parameterizations that may be evaluated.

## METHODS

### Study Design

This analysis used data collected in the ARTnet study of MSM in the United States in 2017–2019. MSM were recruited directly after participating in the American Men’s Internet Study (AMIS) (31), a parent web-based study about MSM sexual health that recruited through banner ads placed on websites or social network applications. At the completion of AMIS, MSM were asked to participate in ARTnet, which focused on sexual network features. ARTnet data collection occurred in two waves (following AMIS): July 2017 to February 2018 and September 2018 to January 2019.

Eligibility criteria for ARTnet were male sex at birth, current male cisgender identity, lifetime history of sexual activity with another man, and age between 15 and 65. Respondents were deduplicated within and across survey waves (based on IP and email addresses), resulting in a final sample of 4904 participants who reported on 16198 sexual partnerships (see Figure 1). The Emory University Institutional Review Board approved the study. Due to the sensitive nature of the data and small sample sizes of participants in some rural geographies, a data use agreement with the ARTnet Principal Investigator (the corresponding author of this paper) will be required before data sharing with external researchers.

**Figure 1.**
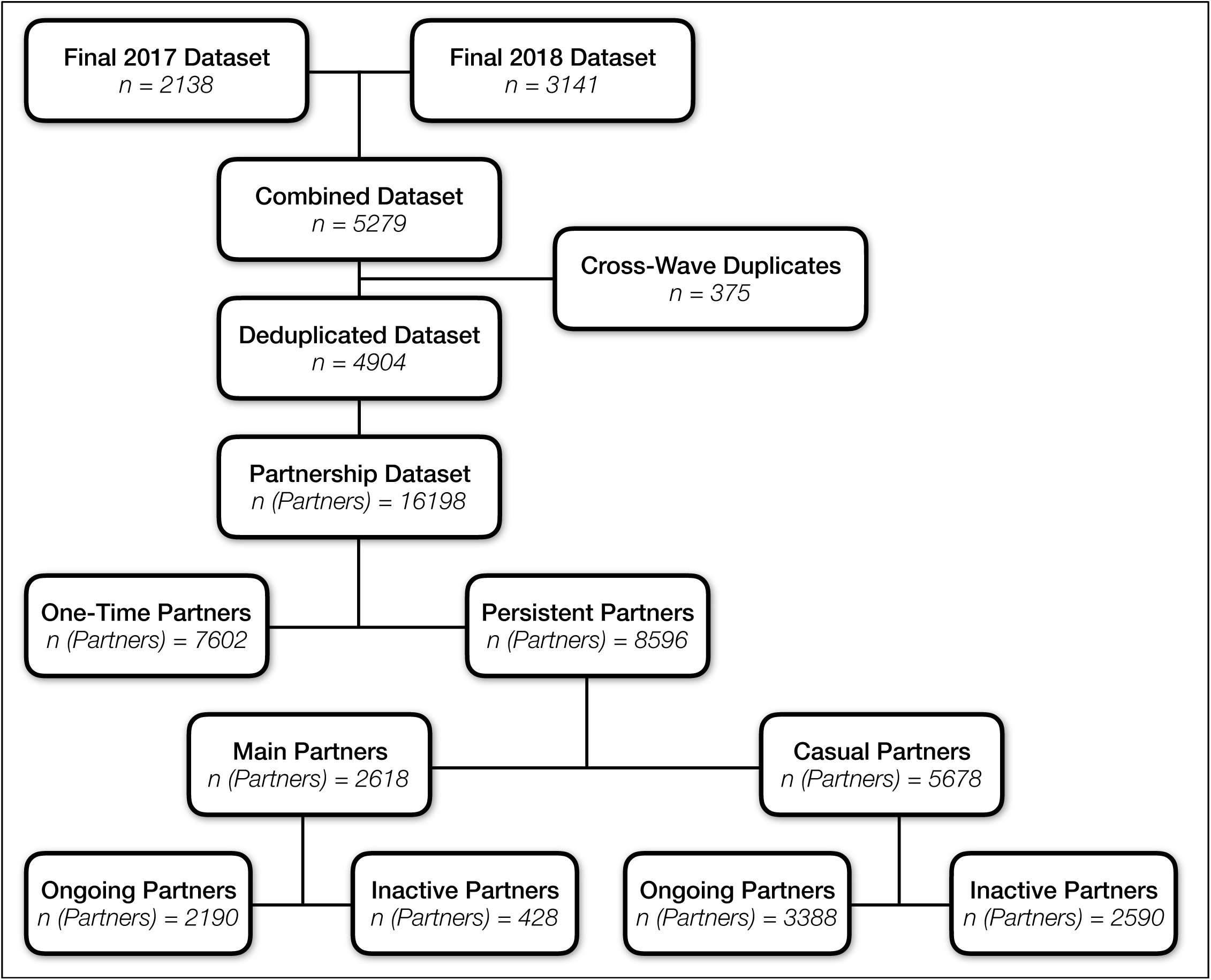
ARTnet Recruitment and Data Type Diagram, with Partnership Stratification by One-Time versus Persistent Partners, Partnership Type, and Ongoing Status of Partnerships.

### Measures

ARTnet participants were first asked about demographic and health-related information. Covariates used in this analysis included race, age, ZIP Code of residence, and current HIV status. ZIP Codes were transformed into Census regions/divisions and urbanicity levels by matching against county databases. Participants reporting as never testing for HIV, having indeterminate test results, or never receiving test results were classified as having an unknown HIV status.

Participants were then asked detailed partner-specific questions for up to most recent 5 partners. The detailed partner-specific questions included attributes of the partner and details about the partnership itself. Partner attributes considered here included age, race/ethnicity, and HIV status. Participants were allowed to report any partner attribute as unknown. When partner age was unknown, age was imputed based on a response to a categorical question (e.g., 5–10 years younger/older, 2–5 years younger/older). Partnerships were classified into three types: “main” (respondent reported they considered this partner a “boyfriend, significant other, or life partner”) casual (someone they have had sex with more than once, but not a main partner), and one-time (17). For one-time partners, we asked for the date that sexual activity occurred. For persistent (main and casual) partnerships, we asked for the date of most recent sex, the date first sex (which could have been prior to the past year), and whether the partnership was ongoing (if the participant expected sexual activity would occur in the future). For each partnership, we asked whether (for one-time) or how frequently (for persistent) oral and anal sex occurred. For this analysis, these questions classified the partnership was oral only, anal only, or both oral and anal.

Outcome measures reported in this analysis include descriptive statistics for characteristics of participants and their reported partnerships, and the aggregate network statistics used to estimate the TERGMs underlying epidemic simulations on dynamic networks. The network statistics include ego degree, attribute mixing in partnerships, and the current length of ongoing partnerships, stratified by the attributes of persons and partnerships. Degree is a property of individuals, whereas mixing and length are properties of partnerships. Degree was defined as the ongoing number of persistent partners measured on the day of the survey (includes main and casual partnerships). Degree is not defined for one-time partnerships, so for these we instead calculated a weekly rate of new contacts by subtracting the total main and casual partners from the total past-year partners, and dividing by 52. Partnership length for main and casual partnerships was calculated by taking the difference between the survey date and the partnership start date. The mean length of ongoing partnerships is the network statistic needed for TERGM estimation; the logic and derivation are explained here (14). Mixing was measured by the relative frequency of partnerships that occurred within and between groups defined by race/ethnicity, age, and HIV status.

### Statistical Analysis

Descriptive analyses of demographic and behavioral characteristics are presented using proportions, means, standard deviations, and medians. Degree was stratified by ego-level characteristics and partnership type, and Poisson regression was used to estimate the mean and 95% confidence intervals. Partnership length was estimated by partnership type for ego-level characteristics and dyadic matching on those characteristics. Age mixing was estimated for both categorical age groups and continuously using absolute differences in age. Mixing was analyzed using joint or row conditional proportions (for categorical attributes), and means and standard deviations for the continuous age difference measure. Analyses were performed in R 3.6.0. The analysis code is available at https://github.com/EpiModel/NetStats.

## RESULTS

Table 1 presents comparative descriptive statistics of deduplicated, unique men who completed the ARTnet study (n = 4904) and the parent AMIS study (n = 21522). ARTnet participants were mainly white non-Hispanic (71.8%), half (52.9%) were aged 15–34, and one-third (36.3%) were from the South. Participants were concentrated in urban counties (Large Central Metro: 43.1%; Large Fringe Metro: 21.5%). Participants reported an HIV prevalence of 8.7%, with 15.3% having an unknown HIV status (mostly due to never HIV testing). Comparing the demographics of ARTnet to AMIS, the race and regional distributions were similar, while ARTnet had more middle-aged participants and more participants from large central metropolitan areas. AMIS had no upper age eligibility restriction, whereas ARTnet had a maximum eligible age of 65. The proportion reporting as HIV-positive was similar, but AMIS had a higher level of unknown HIV status participants (because a higher proportion of AMIS respondents reported never testing for HIV).

**Table 1.**
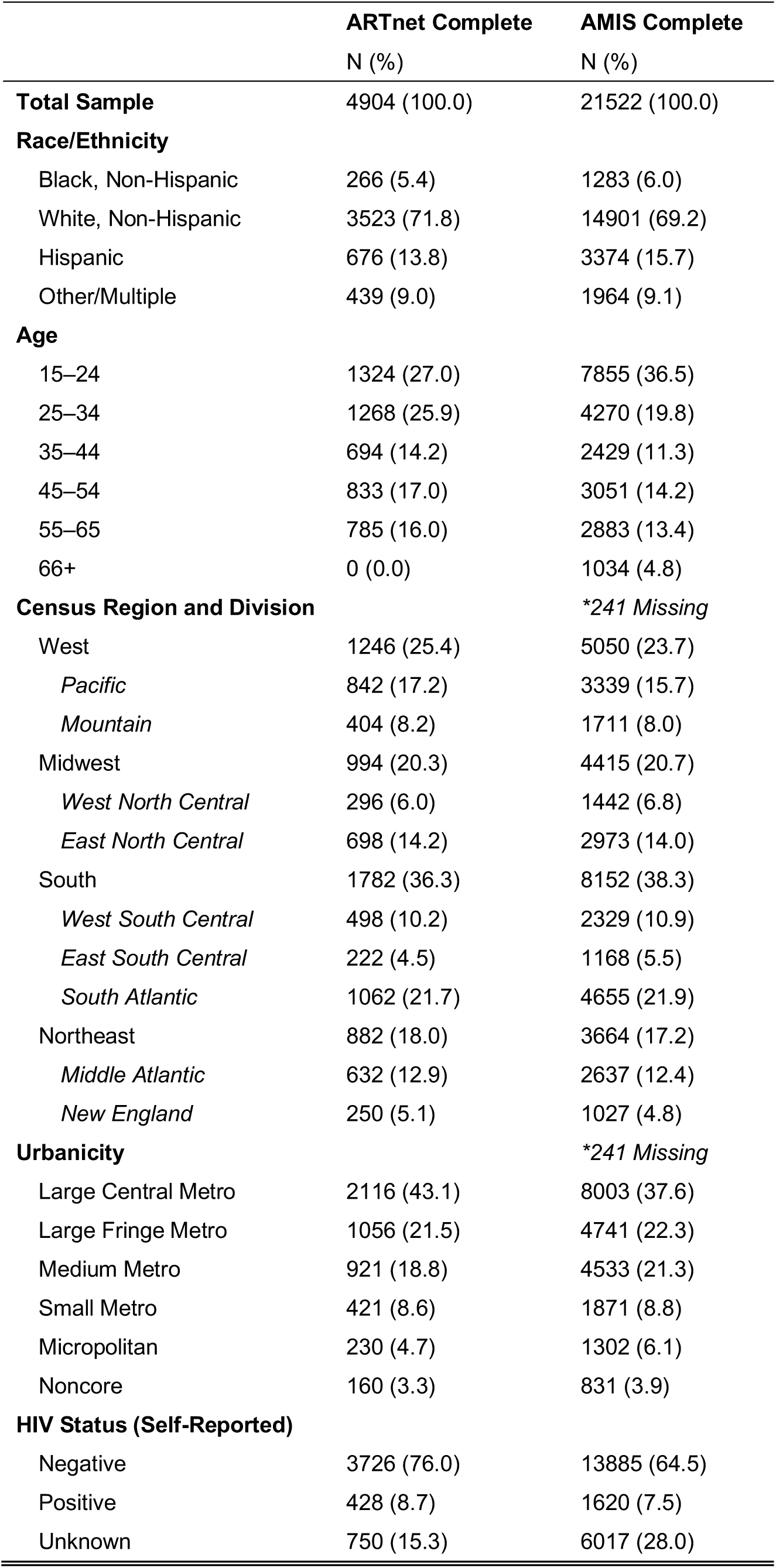
Sample Characteristics of ARTnet Study Participants Compared to and AMIS Study Participants

Over 16000 partnerships were reported in the detailed partnership modules, an average of 3.3 partners per participant (Figure 1). One-time (7602) partners make up almost half of all partnerships reported there, and about twice as many casual partnerships (5978) were reported there than main (2618) partnerships. For the persistent partnerships, the majority (65%) were reported to be ongoing. Main partners were five times more likely to be ongoing than inactive, but they still comprised the minority (about 40%) of all ongoing partnerships.

In Table 2 we move to the first set of network summary statistics: mean degree — the mean number of ongoing persistent partnerships — and weekly rate of one-time partnerships stratified by attributes of participants, overall, and by partnership type (main or casual) for all (oral or anal sex) partnerships. Supplemental Table 1 provides network degree with sexual partnerships separately categorized by any AI and any OI. Participants reported an average of 1.19 (95% CI = 1.16–1.23) ongoing persistent partners, with higher mean degree for casual partnerships (0.74) than main (0.45). By race/ethnicity, white non-Hispanic MSM had the highest persistent partner mean degree (1.22). Black MSM had the lowest main mean degree (0.32), but the highest casual mean degree (0.81).

**Table 2.** Heterogeneity in Mean Degree and Weekly Rates by Demographics and HIV Status, Stratified by Partnership Type, for All Anal or Oral Sex Partnerships among MSM in the United States

|  | Main and Casual | Main | Casual | One-Time |
| --- | --- | --- | --- | --- |
|  | *Mean (95% CI)* | *Mean (95% CI)* | *Mean (95% CI)* | *Mean (95% CI)* |
| **Total** | 1.19 (1.16, 1.23) | 0.45 (0.43, 0.47) | 0.74 (0.72, 0.77) | 0.163 (0.152, 0.175) |
| **Race/Ethnicity** |  |  |  |  |
| Black, Non-Hispanic | 1.12 (1.00, 1.26) | 0.32 (0.25, 0.39) | 0.81 (0.70, 0.92) | 0.117 (0.080, 0.163) |
| White, Non-Hispanic | 1.22 (1.18, 1.26) | 0.46 (0.44, 0.49) | 0.76 (0.73, 0.78) | 0.172 (0.159, 0.186) |
| Hispanic | 1.14 (1.06, 1.22) | 0.45 (0.40, 0.50) | 0.45 (0.63, 0.76) | 0.156 (0.128, 0.187) |
| Other/Multiple | 1.11 (1.01, 1.21) | 0.41 (0.35, 0.47) | 0.70 (0.62, 0.78) | 0.134 (0.102, 0.171) |
| **Age** |  |  |  |  |
| 15–24 | 0.82 (0.77, 0.87) | 0.42 (0.38, 0.45) | 0.40 (0.37, 0.44) | 0.102 (0.086, 0.121) |
| 25–34 | 1.09 (1.03, 1.15) | 0.52 (0.48, 0.56) | 0.57 (0.53, 0.62) | 0.135 (0.116, 0.157) |
| 35–44 | 1.40 (1.31, 1.49) | 0.51 (0.46, 0.56) | 0.89 (0.82, 0.96) | 0.202 (0.170, 0.237) |
| 45–54 | 1.54 (1.46, 1.63) | 0.45 (0.40, 0.50) | 1.09 (1.02, 1.16) | 0.229 (0.198, 0.263) |
| 55–65 | 1.46 (1.37, 1.54) | 0.35 (0.31, 0.39) | 1.11 (1.03, 1.18) | 0.209 (0.179, 0.243) |
| **Census Region and Division** |  |  |  |  |
| West | 1.17 (1.11, 1.23) | 0.43 (0.40, 0.47) | 0.74 (0.69, 0.79) | 0.175 (0.153, 0.200) |
| *Pacific* | 1.29 (1.21, 1.36) | 0.44 (0.40, 0.49) | 0.84 (0.78, 0.91) | 0.195 (0.167, 0.227) |
| *Mountain* | 0.94 (0.85, 1.04) | 0.42 (0.36, 0.49) | 0.52 (0.45, 0.59) | 0.134 (0.101, 0.173) |
| Midwest | 1.18 (1.12, 1.25) | 0.46 (0.42, 0.50) | 0.72 (0.67, 0.78) | 0.154 (0.131, 0.179) |
| *West North Central* | 1.16 (1.04, 1.29) | 0.49 (0.42, 0.58) | 0.67 (0.58, 0.76) | 0.152 (0.111, 0.200) |
| *East North Central* | 1.19 (1.11, 1.28) | 0.45 (0.40, 0.50) | 0.75 (0.68, 0.81) | 0.155 (0.127, 0.186) |
| South | 1.19 (1.14, 1.24) | 0.46 (0.43, 0.49) | 0.73 (0.69, 0.77) | 0.152 (0.134, 0.170) |
| *West South Central* | 1.23 (1.13, 1.33) | 0.47 (0.41, 0.53) | 0.76 (0.69, 0.84) | 0.143 (0.112, 0.179) |
| *East South Central* | 1.19 (1.06, 1.34) | 0.48 (0.40, 0.58) | 0.71 (0.61, 0.83) | 0.162 (0.115, 0.221) |
| *South Atlantic* | 1.17 (1.11, 1.24) | 0.45 (0.41, 0.49) | 0.72 (0.67, 0.78) | 0.153 (0.131, 0.178) |
| Northeast | 1.25 (1.18, 1.32) | 0.44 (0.40, 0.49) | 0.80 (0.75, 0.87) | 0.182 (0.155, 0.211) |
| *Middle Atlantic* | 1.27 (1.18, 1.36) | 0.44 (0.39, 0.50) | 0.83 (0.76, 0.90) | 0.176 (0.146, 0.211) |
| *New England* | 1.20 (1.07, 1.34) | 0.45 (0.37, 0.54) | 0.75 (0.65, 0.86) | 0.194 (0.145, 0.254) |
| **Urbanicity (County)** |  |  |  |  |
| Large Central Metro | 1.26 (1.22, 1.31) | 0.45 (0.43, 0.48) | 0.81 (0.77, 0.85) | 0.196 (0.177, 0.215) |
| Large Fringe Metro | 1.18 (1.12, 1.25) | 0.42 (0.38, 0.46) | 0.76 (0.71, 0.81) | 0.158 (0.135, 0.183) |
| Medium Metro | 1.14 (1.07, 1.21) | 0.48 (0.44, 0.53) | 0.65 (0.60, 0.71) | 0.131 (0.109, 0.155) |
| Small Metro | 1.12 (1.03, 1.23) | 0.46 (0.40, 0.53) | 0.66 (0.59, 0.74) | 0.114 (0.085, 0.149) |
| Micropolitan | 1.06 (0.93, 1.19) | 0.42 (0.34, 0.51) | 0.63 (0.54, 0.74) | 0.132 (0.090, 0.184) |
| Noncore | 1.11 (0.96, 1.28) | 0.40 (0.31, 0.51) | 0.71 (0.59, 0.85) | 0.141 (0.091, 0.208) |
| **HIV Status** |  |  |  |  |
| Negative | 1.24 (1.20, 1.27) | 0.46 (0.44, 0.48) | 0.78 (0.75, 0.81) | 0.172 (0.159, 0.185) |
| Positive | 1.59 (1.47, 1.71) | 0.50 (0.43, 0.57) | 1.09 (1.00, 1.19) | 0.269 (0.223, 0.321) |
| Unknown | 0.75 (0.69, 0.82) | 0.38 (0.34, 0.42) | 0.37 (0.33, 0.42) | 0.063 (0.046, 0.082) |
| **Partnership Concurrency** |  |  |  |  |
| Concurrent (n, %) | 1290 (26.3%) | 95 (1.9%) | 952 (19.4%) | - |

For persistent partnerships, mean degree increased with age, ranging from 0.82 in MSM aged 15– 24 to 1.46 in men aged 55–65. This age trend was driven mostly by the increases in reported ongoing casual partners among older participants. Mean degree for main partners did not differ greatly across age categories, with a maximum of 0.52 among 25–34-year-old MSM and minimum of 0.35 among 55–64-year-old MSM. In contrast, the number of casual partnerships increased substantially with age: 15–24: 0.40; 55–65: 1.11.

MSM in the Middle Atlantic and Pacific census divisions had the highest total degree (1.27 and 1.29, respectively), while men in the Mountain division had the lowest (0.94). MSM in more urban areas had a higher degree (Large Central Metro: 1.26, Large Fringe Metro: 1.18), while MSM in rural areas had the lowest (Micropolitan: 1.06, Noncore: 1.11). HIV-positive men had a higher total (1.59) and casual (1.09) degree than HIV-negative (1.24, 0.78) or status-unknown (0.75, 0.37) MSM. Overall, 1290 (26.3%) MSM reported being in at least two persistent relationships concurrently (degree 2+ for any combination of main and casual partners). Only 1.9% of MSM reported concurrent main partnerships, while 19.4% reported concurrent casual partnerships.

The mean rate of one-time partnership acquisition was 0.163 per week (95% CI = 0.152–0.175), which translates into 8.5 partners per year. The empirical distribution of rates was highly right-skewed, with 25%, 50%, and 75% quantiles of the distribution at 0.000, 0.038, and 0.154 (Supplemental Figure 1). One-time rates were highest among older and HIV-positive MSM. Supplemental Table 2 provides weekly one-time partnership rates with sexual partnerships separately categorized by any AI versus any OI.

Table 3 presents the cross-tabulation of participants’ degree for main partnerships by degree for casual partnerships. Most MSM (79.2%) had a degree of either 0 or 1 across both main and casual partnerships. There was generally a negative correlation between degree in one partnership type and degree in the other, as shown in the marginal summaries. Mean degree for casual partnerships was 0.84, 0.47, and 0.86 for MSM with a main mean degree of 0, 1, and 2, respectively. Although the mean casual degree was highest for MSM with a main degree of 2, this only represented 2% of MSM. Main mean degree was highest (0.53) among MSM with a casual degree of 0, and was consistently lower for MSM with higher casual degrees.

**Table 3.** Mixing Matrix of Participants by Degree for Across Main and Casual Partnership Type, for All Anal or Oral Sex Partnerships among MSM in the United States

|  |  | Casual Degree |  |  |  | Marginal Summary |  |
| --- | --- | --- | --- | --- | --- | --- | --- |
|  |  | 0 | 1 | 2 | 3 | Frequency | Mean Casual Degree |
| Main Degree | 0 | 1432 | 719 | 320 | 338 | 57% | 0.84 |
|  | 1 | 1463 | 271 | 136 | 130 | 41% | 0.47 |
|  | 2 | 48 | 19 | 21 | 7 | 2% | 0.86 |
| Marginal Summary | Frequency | 60% | 21% | 10% | 10% |  |  |
|  | Mean Main Degree | 0.53 | 0.31 | 0.37 | 0.30 |  |  |

Note: Cells implying concurrency (degree > 1) across main and casual partnerships are shaded.

Table 4 presents estimates of the mean length of the 5578 ongoing persistent sexual partnerships by participant characteristics. Partnership lengths were right-skewed, with the mean length exceeding the median length in every case. On average, main partnerships had been ongoing for 272.5 weeks (over 5 years), while casual partnerships were 133.0 weeks old (approximately 2.5 years). The average length of persistent partnerships was longest among white MSM (192.5 weeks) and shortest among Hispanic MSM (158.7 weeks). Hispanic MSM had the shortest ongoing partnerships for both main (219.3 weeks) and casual partners (119.0 weeks); white MSM had the longest ongoing main partnerships (286.1 weeks) and black men had the longest ongoing casual partnerships (154.6 weeks). The mean length of total, main, and casual partnerships increased with participant age, increasing from 71.1 weeks in MSM aged 15–24 to 274.4 weeks in MSM aged 55–65.

**Table 4.** Heterogeneity in Length (in Weeks) of Ongoing Sexual Partnerships by Demographics and HIV Status of Ego Participant, for All Anal or Oral Sex Partnerships among MSM in the United States

|  | Ongoing Partnerships | All | Main | Casual |
| --- | --- | --- | --- | --- |
|  | *N (%)* | *Mean (SD, Med)* | *Mean (SD, Med)* | *Mean (SD, Med)* |
| **Total** | 5837 (100.0) | 185.6 (267.2, 87.9) | 272.5 (336.8, 140.2) | 133.0 (196.5, 70.1) |
| **Race/Ethnicity** |  |  |  |  |
| Black, Non-Hispanic | 297 (5.1) | 186.3 (234.4, 96.1) | 267.9 (286.1, 151.3) | 154.6 (203.0, 82.0) |
| White, Non-Hispanic | 4284 (73.4) | 192.5 (277.5, 90.9) | 286.1 (349.5, 153.1) | 134.9 (201.3, 69.8) |
| Hispanic | 771 (13.2) | 158.7 (231.1, 78.9) | 219.3 (291.7, 103.9) | 119.0 (169.8, 62.6) |
| Other/Multiple | 485 (8.3) | 166.8 (242.5, 82.0) | 241.6 (301.6, 126.9) | 122.6 (186.4, 67.0) |
| **Age** |  |  |  |  |
| 15–24 | 1079 (18.5) | 71.1 (78.8, 46.6) | 74.6 (72.3, 52.2) | 67.4 (84.9, 37.9) |
| 25–34 | 1382 (23.7) | 142.7 (163.4, 83.9) | 195.9 (177.5, 152.7) | 94.8 (132.5, 49.4) |
| 35–44 | 966 (16.5) | 198.7 (239.8, 104.1) | 321.1 (287.2, 248.0) | 128.8 (172.9, 75.6) |
| 45–54 | 1273 (21.8) | 239.9 (312.5, 117.7) | 430.9 (397.3, 287.4) | 160.5 (226.4, 86.0) |
| 55–65 | 1137 (19.5) | 274.4 (379.7, 119.4) | 576.4 (535.5, 376.9) | 179.5 (248.8, 94.1) |
| **HIV Status** |  |  |  |  |
| Negative | 4602 (78.8) | 186.8 (265.4, 90.4) | 281.0 (334.9, 153.0) | 131.2 (193.7, 70.1) |
| Positive | 677 (11.6) | 223.3 (309.3, 100.9) | 359.6 (414.3, 220.6) | 161.6 (222.1, 79.4) |
| Unknown | 558 (9.6) | 130.0 (212.6, 61.1) | 156.3 (242.2, 70.1) | 102.8 (173.2, 50.0) |

HIV-positive MSM had longer persistent partnerships (223.3 weeks) than HIV-negative MSM (186.8 weeks) or MSM with unknown HIV status (130.0 weeks). However, this was strongly confounded by age. The average ages of HIV-negative, HIV-positive, and HIV-unknown MSM were 37.5, 46.7, and 25.9. In a linear regression model of mean length with only HIV status as the predictor (Supplemental Table 4), HIV-positive MSM had total partnership length that were 36.6 weeks longer than HIV-negative MSM. However, in a model with both HIV status and age as predictors, the difference was only 5.0 weeks longer for HIV-positive MSM; age was associated with both HIV status and partnership length.

Table 5 presents the length of ongoing sexual partnerships by partnership-level characteristics. The main comparisons for race/ethnicity, age group, and HIV status group are between partnerships in which the participant and partner have the same attributes compared to partnerships in which individuals have different attributes. For all main and casual partnerships, those in which both partners were black or those in which both partners were white had the longest length (210.1 and 214.2 weeks, respectively), while partnerships between Hispanic MSM were the shortest (141.5 weeks). Black-black casual partnerships were longer than white-white casual partnerships, but the direction was reversed for main partnerships. By participant age, there was a monotonic increase in the length of all, main, and casual partnerships in matched groups, with main and casual partnerships having similar mean lengths for the youngest age category (66.1 and 65.6, respectively), but with main partnership length increasing more substantially across age groups.

**Table 5.** Heterogeneity in Length (in Weeks) of Ongoing Sexual Partnerships by Demographics and HIV Status of Ego Participant and Partner, for All Anal or Oral Sex Partnerships among MSM in the United States

|  | Ongoing Partnerships | All | Main | Casual |
| --- | --- | --- | --- | --- |
|  | *N (%)* | *Mean (SD, Med)* | *Mean (SD, Med)* | *Mean (SD, Med)* |
| **Total** | 5837 (100.0) | 185.6 (267.2, 87.9) | 272.5 (336.8, 140.2) | 133.0 (196.5, 70.1) |
| **Race/Ethnicity** | *\*113 Missing* | *\*113 Missing* | *\*15 Missing* | *\*98 Missing* |
| Black-Black | 147 (2.6) | 210.1 (239.3, 119) | 244.9 (231.0, 169.9) | 196.6 (242.1, 106.4) |
| White-White | 2795 (48.8) | 214.2 (304.6, 97.4) | 313.0 (375.4, 163.5) | 143.1 (214.7, 74.3) |
| Hispanic-Hispanic | 290 (5.1) | 141.5 (190.9, 79.6) | 190.1 (230.6, 105.7) | 104.8 (144.5, 61.3) |
| Other-Other | 88 (1.5) | 160.7 (213.9, 92.4) | 214.9 (260.0, 126.9) | 101.4 (126.4, 70.2) |
| Unmatched | 2404 (42.0) | 160.4 (228.6, 80.8) | 230.4 (289.8, 116.7) | 125.2 (180.7, 66.7) |
| **Age** | *\*48 Missing* | *\*48 Missing* | *\*5 Missing* | *\*43 Missing* |
| 15–24 | 666 (11.5) | 65.9 (71.7, 41.7) | 66.1 (67.6, 45.4) | 65.6 (77.5, 34.6) |
| 25–34 | 813 (14.0) | 149.7 (155.1, 94.8) | 200.0 (160.8, 167.3) | 93.1 (126.6, 49.0) |
| 35–44 | 309 (5.3) | 266.9 (275.7, 164.9) | 395.5 (283.6, 363.5) | 148.7 (207.5, 81.6) |
| 45–54 | 355 (6.1) | 339.0 (370.7, 173.7) | 540.3 (406.0, 440.3) | 221.3 (290.4, 111.7) |
| 55–65 | 217 (3.7) | 443.8 (533.4, 141.7) | 843.2 (587.9, 835.6) | 227.3 (345.4, 95.9) |
| Unmatched | 3429 (59.2) | 177.5 (253.7, 87.7) | 286.9 (343.7, 153.1) | 131.7 (186.5, 71.1) |
| **HIV Status** | *\*10 Missing* | *\*10 Missing* | *\*4 Missing* | *\*6 Missing* |
| Negative-Negative | 3817 (65.5) | 192.5 (271.9, 92.1) | 283.8 (337.9, 153.3) | 130.8 (193.2, 69.7) |
| Positive-Positive | 214 (3.7) | 269.3 (328.8, 143) | 366.5 (379.8, 280.4) | 210.2 (278.8, 87.6) |
| Unknown-Unknown | 202 (3.5) | 136.0 (230.4, 64.2) | 176.2 (310.9, 61.0) | 107.4 (143.5, 65.9) |
| Unmatched | 1594 (27.4) | 163.6 (246.6, 79.9) | 235.9 (322.7, 117.8) | 131.2 (143.5, 65.9) |

Table 6 presents estimates of mixing by race/ethnicity, age, and HIV status. Among all partnerships, most included a white partner (84.6%). Of 2199 partnerships involving a black partner, 369 (16.8%) were with another black partner. Of 4349 partnerships involving a Hispanic partner, 796 (18.3%) were with another Hispanic partner. Of 12911 partnerships involving a white partner, 7094 (54.9%) were with another white partner. Assortative mixing is shown in a matrix format in Supplemental Table 5. Within-race mixing there is expressed as the proportion of egos with a same-race reported partner (row conditional proportions): 45.3% of black, 34.5% of Hispanic, 16.3% of other, and 60.9% of white MSM had partners within their own group.

**Table 6.**
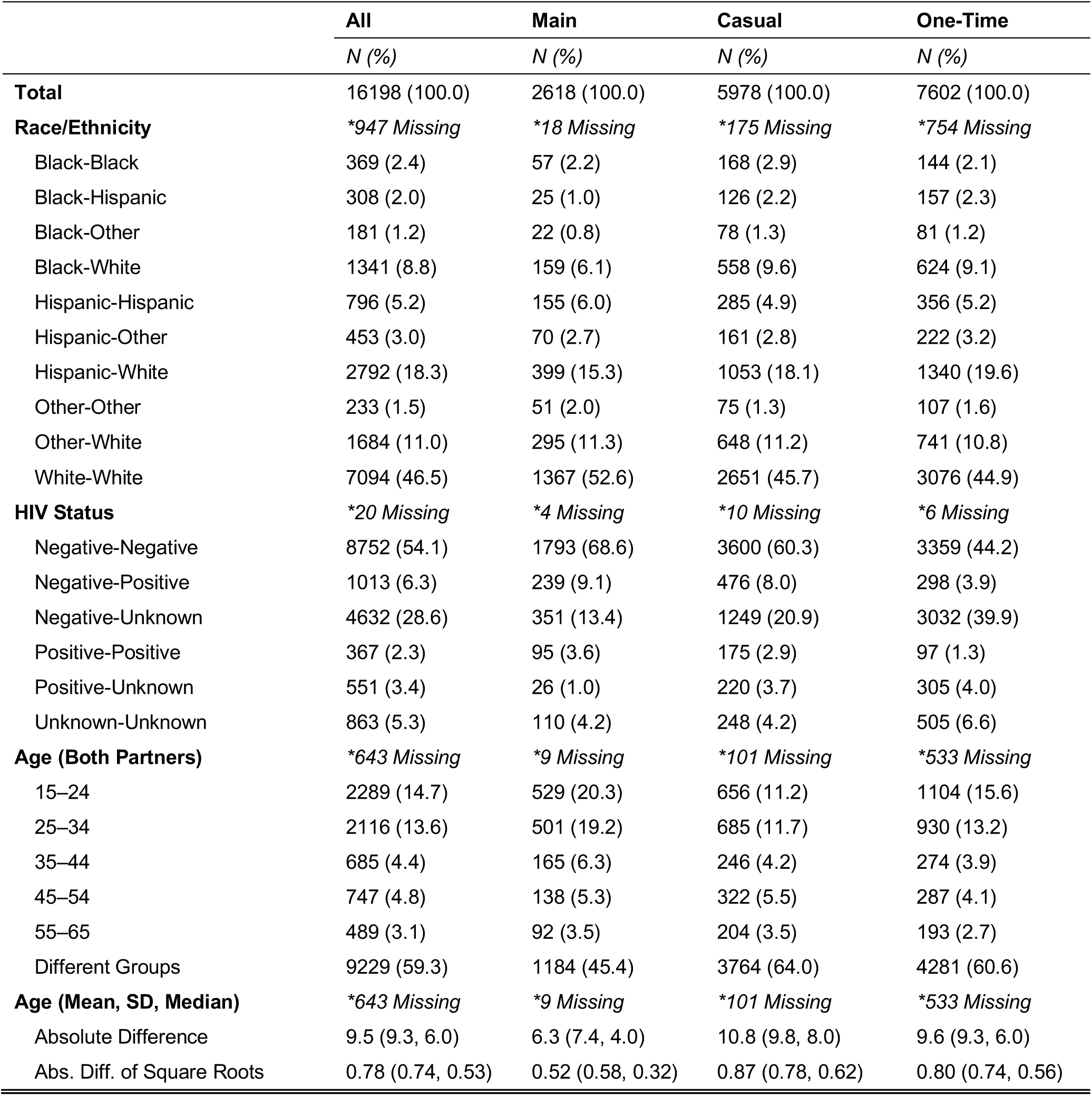
Partnership Mixing by Demographics and HIV Status, Stratified by Partnership Type, for All Anal or Oral Sex Partnerships among MSM in the United States

|  | **All** | **Main** | **Casual** | **One-Time** |
| --- | --- | --- | --- | --- |
|  | *N (%)* | *N (%)* | *N (%)* | *N (%)* |
| **Total** | 16198 (100.0) | 2618 (100.0) | 5978 (100.0) | 7602 (100.0) |
| **Race/Ethnicity** | *\*947 Missing* | *\*18 Missing* | *\*175 Missing* | *\*754 Missing* |
| Black-Black | 369 (2.4) | 57 (2.2) | 168 (2.9) | 144 (2.1) |
| Black-Hispanic | 308 (2.0) | 25 (1.0) | 126 (2.2) | 157 (2.3) |
| Black-Other | 181 (1.2) | 22 (0.8) | 78 (1.3) | 81 (1.2) |
| Black-White | 1341 (8.8) | 159 (6.1) | 558 (9.6) | 624 (9.1) |
| Hispanic-Hispanic | 796 (5.2) | 155 (6.0) | 285 (4.9) | 356 (5.2) |
| Hispanic-Other | 453 (3.0) | 70 (2.7) | 161 (2.8) | 222 (3.2) |
| Hispanic-White | 2792 (18.3) | 399 (15.3) | 1053 (18.1) | 1340 (19.6) |
| Other-Other | 233 (1.5) | 51 (2.0) | 75 (1.3) | 107 (1.6) |
| Other-White | 1684 (11.0) | 295 (11.3) | 648 (11.2) | 741 (10.8) |
| White-White | 7094 (46.5) | 1367 (52.6) | 2651 (45.7) | 3076 (44.9) |
| **HIV Status** | *\*20 Missing* | *\*4 Missing* | *\*10 Missing* | *\*6 Missing* |
| Negative-Negative | 8752 (54.1) | 1793 (68.6) | 3600 (60.3) | 3359 (44.2) |
| Negative-Positive | 1013 (6.3) | 239 (9.1) | 476 (8.0) | 298 (3.9) |
| Negative-Unknown | 4632 (28.6) | 351 (13.4) | 1249 (20.9) | 3032 (39.9) |
| Positive-Positive | 367 (2.3) | 95 (3.6) | 175 (2.9) | 97 (1.3) |
| Positive-Unknown | 551 (3.4) | 26 (1.0) | 220 (3.7) | 305 (4.0) |
| Unknown-Unknown | 863 (5.3) | 110 (4.2) | 248 (4.2) | 505 (6.6) |
| **Age (Both Partners)** | *\*643 Missing* | *\*9 Missing* | *\*101 Missing* | *\*533 Missing* |
| 15–24 | 2289 (14.7) | 529 (20.3) | 656 (11.2) | 1104 (15.6) |
| 25–34 | 2116 (13.6) | 501 (19.2) | 685 (11.7) | 930 (13.2) |
| 35–44 | 685 (4.4) | 165 (6.3) | 246 (4.2) | 274 (3.9) |
| 45–54 | 747 (4.8) | 138 (5.3) | 322 (5.5) | 287 (4.1) |
| 55–65 | 489 (3.1) | 92 (3.5) | 204 (3.5) | 193 (2.7) |
| Different Groups | 9229 (59.3) | 1184 (45.4) | 3764 (64.0) | 4281 (60.6) |
| **Age (Mean, SD, Median)** | *\*643 Missing* | *\*9 Missing* | *\*101 Missing* | *\*533 Missing* |
| Absolute Difference | 9.5 (9.3, 6.0) | 6.3 (7.4, 4.0) | 10.8 (9.8, 8.0) | 9.6 (9.3, 6.0) |
| Abs. Diff. of Square Roots | 0.78 (0.74, 0.53) | 0.52 (0.58, 0.32) | 0.87 (0.78, 0.62) | 0.80 (0.74, 0.56) |

By HIV status, most relationships (89.0%) involved an HIV-negative partner (Table 6). Of 14397 partnerships involving a HIV-negative partner, 8752 (60.8%) were with another HIV-negative partner. Of 1931 partnerships involving a HIV-positive partner, 367 (19.0%) were with another HIV-positive partner, while 551 (28.5%) were with a partner of unknown status. The proportion of relationships with a partner of unknown HIV status was lowest within main partnerships (487, 18.6%), higher among casual partnerships (1717, 28.7%), and greatest within one-time partnerships (3842, 54.4%). Supplemental Table 6 presents a mixing matrix version of these results.

Finally, by age, 59.3% of partnerships were among partners in different age categories, ranging from 45.4% among main partnerships to 64.0% among casual partnerships. Across all partnerships, the mean difference in age between ego and alter was 9.5 years, with main partners only differing by 6.3 years on average compared to an average of 10.8 years among casual partners. The absolute age differences in one-time partnerships was slightly lower than those in casual partnerships.

## DISCUSSION

In this paper, we present the first detailed egocentric network statistics of MSM from across the United States. This analysis highlighted three network features critical for driving the prevalence and dynamics of HIV/STI epidemics in this target population: network degree, assortative mixing, and partnership length. This analysis suggests that MSM across geographies and demographics may be at sustained risk for HIV/STI acquisition and transmission through a combination of both high cumulative numbers of sexual partners and high ongoing mean degree among persistent partnerships. The reported partnerships involve high levels of disassortative mixing across HIV statuses, and between race and age groups with large preexisting differences in HIV/STI prevalence. Overall, these network descriptive statistics provide a foundation for understanding the latest trends in HIV/STI epidemiology among U.S. MSM, to be supplemented by future statistical and mathematical modeling to investigate the population-level implications for disease transmission dynamics and prevention strategies.

HIV/STI risk in MSM sexual networks are influenced by degree, mixing, and duration in combination (14). The past decade has seen a stabilization of HIV incidence despite expanding access to and use of highly effective HIV prevention tools (32), and an increase in three major bacterial STIs (gonorrhea, chlamydia, and syphilis) despite the availability of antibiotics (33). Recent national analyses have shown a broad increase in total and casual past-year partners among MSM that could explain these disease trends (34). However, cumulative statistics like total past-year partnerships are not fully informative of current and future epidemic potential due to the offsetting effects of degree and duration in dynamic partnership networks (35). Lower mean degree (such as that observed among younger MSM in our study) does not necessarily translate to lower risk. Because of their shorter partnership durations, younger MSM have higher partnership turnover rates than older MSM. Dynamic network statistics such as the “forward reachable path” (36) and dynamic network-based mechanistic models (14) are needed to disentangle degree and duration to fully understand these results.

These findings also contribute to our understanding of HIV racial disparities. HIV incidence has remained stable among black MSM, who account for the greatest number of new HIV diagnoses, and increased 12% among Hispanic MSM despite decreasing 14% among white MSM (5). A long-standing puzzle has been to explain the causes of these disparities in light of evidence finding equal or lower risk levels for the race groups (black MSM) with the highest incidence (17,37,38). One hypothesis has been assortative mixing by race: that black MSM are at greater risk for HIV because they are exposed to HIV at a higher rate because most of their contacts are with black MSM in whom prevalence is higher (39). While assortative mixing may result in higher HIV exposure cross-sectionally, modeling suggests assortative mixing alone cannot generate or sustain disparities without differences in risk between groups (40). A combination of preferential mixing plus risk-amplifying network features (e.g., higher mean degree) would be needed to explain the disparity over time. In our study, we found minor differences in mean degree by race/ethnicity, with the fewest ongoing main and casual partnerships among black and Hispanic men, respectively, and a white/black difference of nearly 3 one-time partners per year. By this measure alone, network risks were lower among black MSM, consistent with previous research (41,42). However, we found other network features that could be contributing to these disparities. Black MSM consistently had longer casual partnerships compared to white MSM, lengthing the opportunities for disease exposure.

We found some evidence for serosorting (mixing by known HIV status), as concordant status relationships made up more than 20% of total relationships of HIV-positive respondents, despite HIV prevalence in the population of less than 9%. This is not necessarily a sign of serosorting (preferential mixing in partnership formation), however, because the homophily may also reflect HIV transmissions that occur within a partnership that started as discordant (43). Notably, the proportion of partnerships involving an unknown partner was higher among casual and one-time partnerships, making up more than half of all one-time partnerships.

One strength of the ARTnet survey is that it included MSM across a wide age range (15 to 65). We found that HIV-positive respondents reported the longest partnerships, but this was strongly confounded by age. As an incurable infection, HIV prevalence increases with age (44,45), but younger MSM account for most new HIV diagnoses (44). Therefore, explanatory factors for increased risk acquisition among younger MSM are needed. Age was associated with an increase in casual and one-time partners, with the rate of partner acquisition among 35 to 65-year-old MSM twice that of those aged 15 to 24 years.

There was a strong association between age and the duration of main partnerships, with a weaker association for casual partners. Younger MSM in our study may have reported lower degree, but the relatively weak age-based homophily and shorter partnerships could result in greater partner turnover with and increased risk of HIV exposure through contacts with older partners.

By geography and urbanicity, we found small differences in network risk that do not reflect the underlying spatial distribution of HIV burden. The distribution of HIV diagnoses (52% in 2017) and HIV prevalence (46% in 2015) is greatest in the South region, but this is also confounded by race/ethnicity (5). We did not observe higher mean degree of persistent partners or a higher rate of one-time partners in the South. Men from the Pacific and Middle Atlantic divisions had the greatest number of ongoing partnerships, driven by more ongoing casual partnerships. These are also regions where access to and use of prevention tools (such as PrEP) are the highest (46). This may reflect two fundamental changes in the epidemiological context for MSM over the past decade. First, in the era of pharmacological elimination of HIV acquisition and transmission risk (47), the spatial distribution of HIV may be driven more by heterogeneity in the use of prevention tools rather than differences behavioral risk (48). Second, the greater availability of prevention tools in some areas may cause an increase in behavioral risk through the knowledge that HIV biological risk is low or eliminated (49).

Finally, with respect to urbanicity, differences in population size, as might be observed in urban areas compared to rural areas, change the ratio of edges (partnerships) to nodes (individuals). This means that either the density of the network or the mean degree will differ in networks (or subnetworks) of different sizes (14). Some studies have interpreted differences in network density, by itself and without consideration to network size, to have meaning in the HIV/STI context (41). However, HIV/STI modeling studies typically assume frequency dependence, which preserves mean degree over network density when network size fluctuates (50,51). In this study, however, we found some evidence of a correlation between mean degree and population size of MSM, primarily with respect to differences in casual mean degree and one-time partnership rates (8–10 in Large Central and Large Fringe Metros versus 6–7 in less urban counties). For meta-population models, such as those that consider migration between rural and urban areas (52), the frequency dependency assumption and corresponding preservation of mean degree may not be warranted.

### Limitations

There are some limitations to this analysis. As a self-reported survey, there are concerns about possible recall and self-report biases associated with answering questions about sexual behavior, including reporting detailed information on all partners in the prior year or the length of a partnership. Certain partner information necessary for this analysis, such as HIV status, is influenced by disclosure and may not have been known to survey respondents. However, in most cases we allowed a “don’t know” or “prefer not to answer” survey response. Additionally, the recruitment method resulted in a convenience sample of MSM that may not be representative of all MSM, particularly racial/ethnic minority MSM who may be under-represented in online studies (53,54). In the U.S., there are no population-based probability samples of MSM, with the largest ongoing study (the National HIV Behavioral Surveillance study) relying on venue-based recruitment that does not sample MSM who do not frequent gay-oriented social venues (34). However, just as there are methods for weighting venue-based data (55), our web-based ARTnet data may be weighted to account for biases in sampling. We did not weight currently as a first descriptive analysis of this dataset, but future localized analyses of ARTnet can and should weight.

### Conclusions

Estimates of network summary statistics among MSM that are up to date and broad in both demographic and geographic scopes, such as ARTnet, are essential. They provide the necessary empirical foundation for understanding population level HIV/STI transmission dynamics, as individual-level risks alone do not explain the size or heterogeneity in HIV/STI burden among MSM in the U.S. (34,56). ARTnet demonstrates the feasibility of collecting these data quickly and nationally. Unlike other forms of network data collection, this type of egocentric sampling can be used for routine public health monitoring purposes. These network statistics do not tell the full epidemiological story, because they interact with the many factors that determine engagement in HIV-related prevention and care to generate epidemic dynamics. Mathematical modelling, and specifically network-based modeling that incorporates principled statistical estimation from egocentrically sampled network data, is needed to fully represent this transmission system. The descriptive statistics presented here are a first but important step in the process. These study designs and data, combined with the software tools for network model estimation and simulation (14), provide a promising new framework for investigating and addressing, the critical challenges in HIV/STI prevention science.

## Data Availability

Original analysis code is available and data is available with a data sharing agreement with the study Principal Investigator (Dr. Jenness)

https://github.com/EpiModel/NetStats

